# High burden of anti-microbial resistance among neonatal blood stream infections in Southeast Asia: results of the NeoSEAP study

**DOI:** 10.1101/2023.09.11.23295383

**Authors:** Benjamin F R Dickson, Nina Dwi Putri, Riyadi Adrizain, Leny Kartina, Maria Esterlita Villaneuva Uy, Gayana Gunaratna, Chau Le, Hoang Tran, Hương Nguyễn Xuân, Distayay Sukarja, Tetty Yuniati, Martono Utomo, Nguyen Thi Kieu Trinh, Hoang Nguyen Thanh Thuy, Tran Thi Cam Tu, Le Tuyet Hong, Siew Moy Fong, Michelle Harrison, Phoebe C M Williams, the NeoSEAP consortium

## Abstract

**Background:** Progress on neonatal sepsis has remained modest in recent decades and is threatened by the global rise of antimicrobial resistance. The Southeast Asian region has a high burden of both neonatal sepsis and antimicrobial resistance. Despite this, their remains a lack of robust epidemiological data on the causes of neonatal sepsis and the prevalence of AMR in the region.

**Methods:** We evaluated the causes of neonatal sepsis and AMR burden in 10 sites across five countries in South and Southeast Asia (Sri Lanka, Indonesia, The Philippines, Malaysia and Vietnam). Retrospective data on all blood cultures collected from neonates between 1^st^ January 2019 – 31^st^ December 2020 were extracted from laboratory records. Data were also collected on the availability of clinical resources, the implementation of infection prevention and control strategies, and antimicrobial prescribing practices.

**Findings:** A total of 1,528 blood cultures were positive for significant isolates over the study period. Gram-negative pathogens predominated (1,163/1,528, 76.1%) with the most frequently isolated pathogens *Klebsiella* spp. (408/1,528, 26.7%) and *Acinetobacter* spp. (261/1,528, 17.08%) Among Gram-negative Enterobacteriaceae pooled resistance to ampicillin, gentamicin, third-generation cephalosporins (ceftriaxone and/or cefotaxime) and carbapenems was 75% (193/257), 59% (393/665), 67% (441/655) and 18.6% (125/672). For Gram negative non-Enterobacteriaceae resistance to gentamicin and carbapenems was 76.6% (326/282) and 69.7% (207/297).

**Interpretation:** Neonatal sepsis among study sites was caused predominantly by Gram-negative pathogens and associated with high levels of non-susceptibility to common empirical treatment regimes.

## Introduction

Significant progress has been made in reducing child mortality in recent decades[1]. Despite this, neonatal deaths have only witnessed relatively modest improvements[1,2]. Sepsis remains an important cause of neonatal mortality with an estimated half a million deaths occurring annually[2]. Many survivors also face long-term sequelae including higher post-discharge mortality rates, cognitive and physical disabilities[3]. This burden disproportionally affects low- and middle-income countries (LMICs), where 85% of neonatal sepsis-related deaths occur[4].

Concurrently, the rise of antimicrobial resistance (AMR) threatens to halt, or even reverse recent global health gains[5]. Deaths from AMR are projected to outpace those from diabetes and cancer combined by 2050[5]. The AMR burden already does, and will continue to, inequitably fall on LMICs, where fragile health systems are less able to front this emerging challenge[5].

Neonates are particularly vulnerable to the rise of AMR. The rapid progression and non-specific presentation of neonatal sepsis drives high-rates of antimicrobial use. Simultaneously, premature infants experience long hospital admissions, which increase the probability of AMR colonisation and nosocomial infections. This risk is compounded in LMICs, where sepsis is predominantly caused by gram-negative infections, and over-burdened health systems foster horizontal AMR transmission[6]. Emerging data in Africa and South Asia have demonstrated concerning levels of resistance to common first-, second- and third-line empirical treatment regimes to the extent that it is now estimated that one-third of neonatal sepsis deaths are directly attributable to AMR [7-11].

The Asian region has the highest global population, accounts for 40% of global cases of neonatal sepsis, and is anticipated to have the greatest number of deaths attributable to AMR by 2050[5,12,13]. Despite this, there remains a paucity of data on both the causes neonatal sepsis and the burden of AMR in this region [14]. These data are particularly critical in neonatal sepsis, where rates of culture-negative sepsis are high, so treatment relies heavily on empirical regimes based on known pathogen distributions, and susceptibility profiles[15].

This study therefore sought to contribute to this epidemiological gap by evaluating the causes of neonatal sepsis, and the associated burden of antimicrobial resistance across several countries in South and Southeast Asia. To provide context to these data, it also assessed available healthcare resources, the implementation of infection prevention and control (IPC) strategies, and antimicrobial prescribing practices at each site.

## Methods

### Study design

This project was conducted by the Neonatal Sepsis in Southeast Asia and the Pacific (NeoSEAP) consortium. It was a multicentre, observational study conducted in 10 sites across five countries in South and Southeast Asia (Sri Lanka, Indonesia, The Philippines, Vietnam and Malaysia). The study included three components: 1) a survey of available healthcare resources and utilisation of IPC strategies; 2) a retrospective audit of blood cultures collected from neonates over a 24-month period, and 3) a prospective antimicrobial point prevalence survey (PPS) evaluating the treatment, demographic and clinical data of all infants less than 180 days old.

### Site selection

The study included countries with identified gaps in epidemiological data on the causative pathogens of neonatal sepsis and their associated AMR burden. Sites within each country were then selected through active recruitment.

### Data collection

Data were collected using the electronic data-collection instrument REDCap (www.redcap.org). Information on the availability of hospital resources and the implementation of IPC strategies was obtained from hospital records and staff, and then verified by site-visits were possible. During the PPS, all admitted infants receiving antimicrobial therapy at 8am on a pre-defined date between December 2022 and January 2023 were identified. Demographic, clinical and treatment data were then extracted from hospital records by clinical staff.

Microbiological data from all blood cultures collected from neonates (defined as ≤ 28 days corrected gestational age) from 1^st^ January 2019 to 31^st^ December 2020 were systemically extracted from electronic and/or paper laboratory records. Data on the result, species (from a pre-defined list) and antimicrobial susceptibility profile were recorded. Positive cultures from the same patient with the same organism within four weeks were considered a repeat and were excluded from the study.

The indication for neonatal blood culture collection in all sites was clinical instability and/or suspicion of a severe infection. Blood cultures were processed with the BacTec FX System (Becton Dickinson, Sparks USA) and/or BacT/ALERT 3D (bioMerieux, Inc. Durham USA) automated microbial detection systems. Positive cultures underwent identification and antimicrobial susceptibility testing using VITEK-2 (bioMerieux, Inc. Durham, USA), Phoenix (Becton Dickinson, Sparks USA) automated analysers, and conventional methods. In Sri Lanka, no automated analyser is available with pathogen identification for gram-negative pathogens limited to lactose-fermentation status, rather than genus or species level due to resource constraints. Antimicrobial susceptibility results were interpreted according to breakpoints established by Clinical and Laboratory Standards Institute (CLSI). Intermediate results were classified as resistant for the analysis.

### Data analysis

Data were managed in Microsoft Excel (Version 16) and analysed in Stata 16 (Stata corporation, TX). Proportions with exact 95% binomial confidence intervals were generated for pathogen frequency and AMR prevalence and compared using Chi-squared or Fisher exact tests. For the three Indonesian sites, country-level pooled coverage estimates for Ampicillin plus gentamicin, non-pseudomonal active third generation cephalosporins and carbapenems were generated using a weighted-incidence syndrome combination antibiogram (WISCA) as previously described[16]. No formal statistical evaluations of the association between AMR prevalence, resource availability and infection prevention and control strategies were undertaken because of the limited number of sites. The analysis of the PPS and fungal isolates will be reported separately.

### Ethical approval

Ethics approval was obtained from Universitas Indonesia, Jakarta; Institutional research board, Dr. Soetomo Hospital, Surabaya (0339/KEPK/XII/2021); and Komite Etik Penelitian RSUP Dr. Hasan Sadikin, Bandung. Because the study was clinical audit, a waiver of participant consent was approved by the HREC committees at each site, and a waiver of ethical approval was granted for the Tam Tri Hospital Network.

## Results

### Site characteristics and resources

The characteristics and location of the 10 study sites included in the study are outlined in Table 1 and Figure 1. Seven of the sites were public tertiary teaching hospitals, whilst the remaining three were private referral hospitals (all located in Vietnam). All hospitals delivered infants on-site, with a median of 1,594.5 (IQR: 1,308 – 4,918) births per year. The median bed capacity (both post-natal ward and admitted) was 87 (IQR: 46 – 120) with an average of 1,168.5 (IQR: 247 – 1895) admissions per year. Among admitted infants the average proportion of premature infants was 41.9% (IQR: 11.3 – 51.8%). All ten sites had access to total-parenteral nutrition whilst nine (90%) were equipped with incubators, eight (80%) offered invasive ventilation and four (40%) had established donor milk programs.

**Table 1.** Characteristics of NeoSEAP sites.

| Table 1. Characteristics of NeoSEAP sites |  |  |  |  |  |  |  |  |  |  |  |
| --- | --- | --- | --- | --- | --- | --- | --- | --- | --- | --- | --- |
|  |  | Indonesia |  |  | Philippines | Sri Lanka | Malaysia | Vietnam |  |  |  |
| Site |  | 1 | 2 | 3 | 4 | 5 | 6 | 7 | 8 | 9 | 10 |
| Care type | Ownership | Public Tertiary | Public Tertiary | Public Tertiary | Public Tertiary | Public Tertiary | Public Tertiary | Public Tertiary | Private Referral | Private Referral | Private Referral |
| Beds | Post-natal ward | 2 | 24 | 28 | 48 | 30 | 104 | 107 | 25 | 40 | 40 |
|  | Special-care Nursery | 17 | 24 | 40 | 30 | 6 | 58 | 12 | 10^ | 54^ | 6 |
|  | HDU | 19 | 44 | 32 | 0 | 10 | 24 | 111 |  |  | 0 |
|  | ICU | 30 | 14 | 20 | 30 | 8 | 18 | 49 |  |  | 0 |
|  | Total | 68 | 106 | 120 | 108 | 54 | 204 | 279 | 25 | 40 | 46 |
| Staffing | Nurses (Total FTE) | 116 | 64 | 53 | 42 | 25 | 162 | 61 | 3 | 0 | 12 |
|  | Doctor (Total FTE) | 14 | 7 | 8 | 24 | 12 | 25 | 14 | 1 | 0 | 4 |
|  | Consultant (Total FTE) | 1 | 3 | 4 | 7 | 2 | 1 | 3 | 1 | 0 | 1 |
| Deliveries | Deliveries on-site | Yes | Yes | Yes | Yes | Yes | Yes | Yes | Yes | Yes | Yes |
|  | Birth per year | 1,508 | 1,681 | 853 | 3,365 | 4,918 | 14,052 | 10,159 | 492 | 1,308 | 1,339 |
| Admissions | Total | 1,433 | 1,895 | 1,490 | 904 | 398 | 6,081 | 2,448 | 52 | 215 | 247 |
|  | Premature | 823 | 797 | 715 | 864 | 206 | 986 | 1,023 | 1 | 12 | 28 |
| Services | Neonatal Surgery | Yes | Yes | Yes | Yes | No | Yes | Yes | Yes | No | No |
|  | Incubators (number) | 17 | 40 | 43 | 27 | 16 | 29 | 17 | 1 | 0 | 1 |
|  | Non-invasive ventilation | Yes | Yes | Yes | Yes | Yes | Yes | Yes | Yes | No | Yes |
|  | Invasive ventilation | Yes | Yes | Yes | Yes | Yes | Yes | Yes | Yes | No | No |
|  | Inotropes | Yes | Yes | Yes | Yes | Yes | Yes | Yes | No | Yes | Yes |
|  | TPN | Yes | Yes | Yes | Yes | Yes | Yes | Yes | Yes | Yes | Yes |
|  | Donor-milk | Yes | No | No | Yes | No | No | Yes | No | No | Yes |

^Neonatal beds on paediatric ward

**Figure 1.**
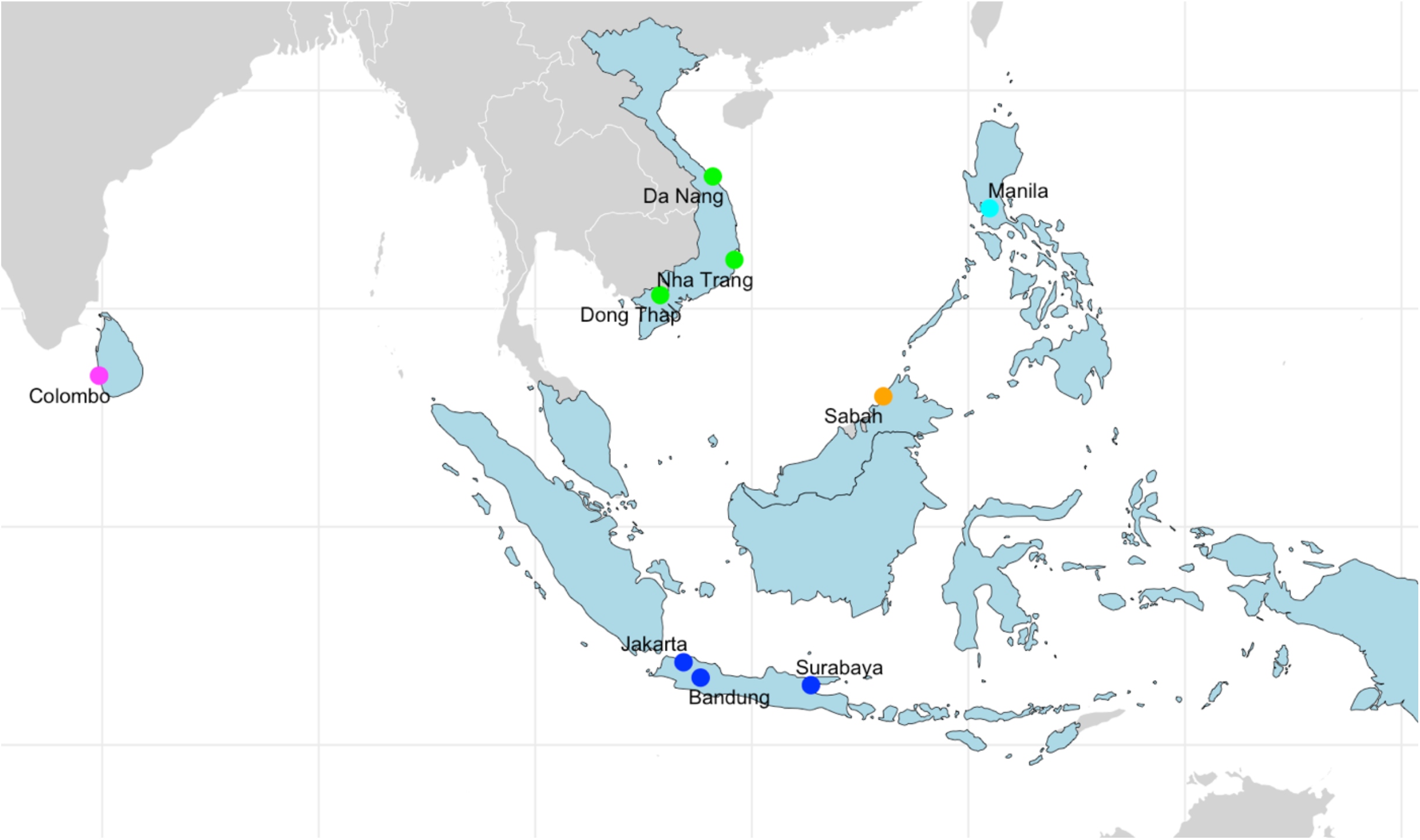
Map of included countries and study sites

### Infection prevention & control strategies

Figure 2. shows a heatmap of the infection prevention and control resources available and strategies implemented at each site. All ten sites had established local IPC guidelines, and the majority (90%) employed IPC specific staff. Antiseptic hand rub and gloves were always available at 6 (60%) and 9 (90%) of sites respectively. The median proportion of infants breastfeeding on discharge was 80% (IQR 70 – 92%). Kangaroo care was routine practiced by 9 (90%) sites.

**Figure 2.**
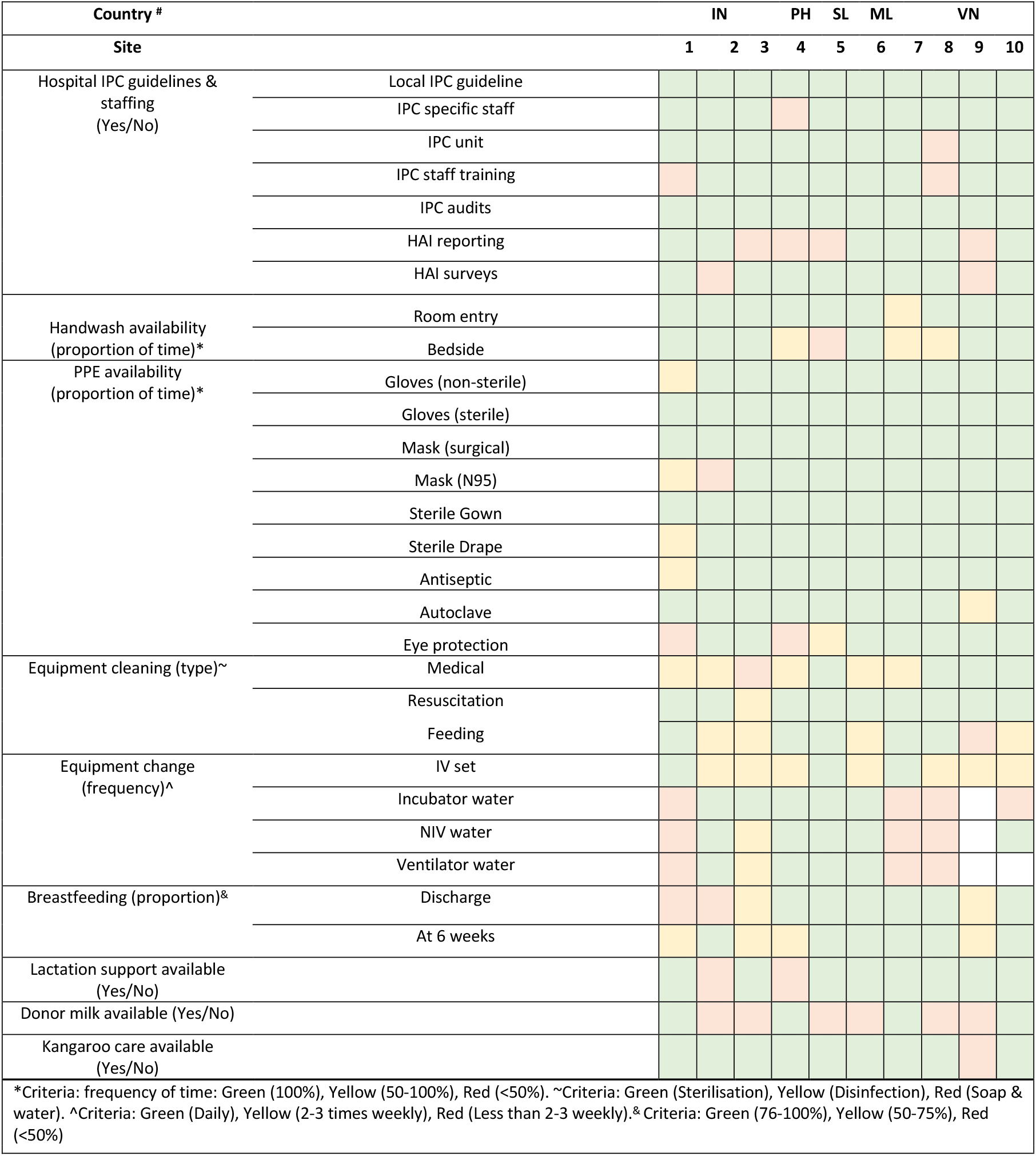
Heatmap of resource availability and Infection prevention & control strategies by site.

### Blood culture collection and empirical treatment regimes

Table 2. outlines the blood culture collection practices and empirical treatment regimens by site. Seven of the 10 sites (70%) routinely collected blood-cultures prior to starting antibiotics, with 48 hours most frequently considered the cut-off for a negative culture (range: 48 hours – five days).

**Table 2.** Empirical antimicrobial recommendations by site.

| Country |  | Indonesia |  |  | Philippines | Sri Lanka | Malaysia | Vietnam |  |  |  |
| --- | --- | --- | --- | --- | --- | --- | --- | --- | --- | --- | --- |
| Site |  | 1 | 2 | 3 | 4 | 5 | 6 | 7 | 8 | 9 | 10 |
| Blood cultures | Routine pre-treatment | Yes | Yes | Yes | Yes | Yes | Yes | Yes | No | No | No |
|  | Culture negative cut-off (time) | 5 days | 48 hours | 72 hours | 48 hours | 48 hours | 48 hours | 72 hours | 48 hours | 5 days | 48 hours |
| Treatment guidelines used (update year) |  | Local (2019), National (2019), WHO | Local (2020), National (2018), WHO | Local (2016), National (2016), WHO | Local (2016), National (2020) | Local (2016), National (2014), NICE | Local (2020), National (2018), WHO | Local (2021), National (2016), WHO | National (2022) | Local (2020), National (2015), Children's Hospital 1 | Local (2020), National (2015), WHO, NICE |
| Empirical antimicrobial regimes* | EONS | AMP 50 mg/kg TDS<br>GEN 5 mg/kg OD | AMP 100 mg/kg BD<br>GEN 4 mg/kg OD | AMP 50 mg/kg BD<br>GEN 5 mg/kg OD | AMP 50 mg/kg TDS<br>AMK 15 mg/kg OD | PEN 100 mg/kg BD<br>GEN 5 mg/kg OD | AMP 50 mg/kg BD<br>GEN 5 mg/kg OD | AMP 50 mg/kg TDS<br>GEN 4 mg/kg OD | AMP 150 mg/kg BD<br>GEN 5 mg/kg OD | AMP 150 mg/kg TDS<br>AMK 15 mg/kg OD | AMP 150 mg/kg TDS<br>GEN 4 mg/kg OD |
|  |  |  |  |  |  |  |  |  |  | CTX 100 mg/kg BD |  |
|  | LONS | AMP/Sulbac. 50 mg/kg TDS<br>GEN 5 mg/kg OD | CTX 150 mg/kg TDS<br>. . | AMP 50 mg/kg BD<br>GEN 5 mg/kg OD | CAZ 100 mg/kg TDS<br>AMK 15 mg/kg OD | PEN 100 mg/kg BD<br>CTX 50 mg/kg TDS | Cloxacillin 50mg/kg BD<br>GEN 5 mg/kg OD | Ticarcillin 80 mg/kg QID<br>AMK 15 mg/kg OD | CTX 150 mg/kg TDS<br>GEN 5 mg/kg OD | AMP 150 mg/kg TDS<br>CTX 150 mg/kg TDS | AMP 150 mg/kg BD<br>GEN 4 mg/kg OD |
|  |  |  |  |  |  |  |  |  |  | AMK 15 mg/kg OD |  |
|  | Meningitis | AMP/Sulbac. 50 mg/kg TDS<br>GEN 5 mg/kg OD | MEM 40 mg/kg TDS | CTX 100 mg/kg BD<br>GEN 8 mg/kg OD | AMP 50 microg/kg TDS<br>AMK 15 mg/kg OD | CTX 50 mg/kg TDS | PEN 100,000 IU/kg BD<br>CTX 50 mg/kg BD | AMP 50 mg/kg QID<br>GEN 4mg/kg OD | CTX 100 mg/kg OD<br>GEN 5mg/kg OD | AMP 300 mg/kg QID<br>AMK 15 mg/kg OD | AMP 300 mg/kg TDS<br>GEN 4mg/kg OD |
|  |  |  |  |  |  |  |  | CTX 50mg/kg QID |  | CTX 200 mg/kg QID | CTX 300 mg/kgTDS |
| Anti-fungal prophylaxis | Recommended | No | No | Nystatin 100,000 IU PO TDS^ | Fluconazole 3mg/kg IV q72h~ | No | No | No | No | No | No |

\*Colour refers AWARe classification: Access (green), Watch (yellow), Reserve (red). All antibiotic doses are intravenous. OD (Once daily), BD (Twice daily), TDS (Three times daily), QID (Four times daily).
^Nystatin criteria: Weight <1500g; GA <32 weeks. ~Fluconazole criteria: Weight <= 1500g + broad spectrum Abx;
Weight >1500g + NBM >5 days. AMP (Ampicillin), GEN (Gentamicin), AMK (Amikacin), MEM (Meropenem), CAZ (Ceftazidime), CTX (Cefotaxime or Ceftriaxone)

Most sites (90%) used multiple sources to inform empirical management practices of neonatal early- and late-onset sepsis and meningitis including national (100%), local (90%), World Health Organization (60%) and United Kingdom National Institute for Clinical Evidence (NICE) (20%) recommendations. Most local and national guidelines (89%) were published at least two or more years prior to the study. The predominant regime used for early-onset sepsis was ampicillin plus gentamicin (70%). In contrast, regimes for late-onset sepsis and meningitis were more varied, but most containing an aminoglycoside plus a penicillin-derivative and/or cephalosporin. Antibiotics classified in the ‘Watch’ group by the World Health Organization (WHO), were incorporated into empirical treatment regimens for late-onset sepsis, and meningitis in 60% and 80% sites respectively. Two sites (20%) recommended anti-fungal prophylaxis for at-risk infants: one using nystatin and the other fluconazole.

### Neonatal blood cultures

The number of neonatal blood cultures and the species isolated are shown in Table 3. Over the study period, there were a total of 1,528 positive blood cultures with clinically significant isolates as outlined alongside denominator data in Table 4. The median blood culture positivity rate from sites with available denominator data was 11.21 (IQR: 7.9 – 21.04%). Among significant isolates, gram-negatives predominated (1,163/1,528, 76.1%), followed by gram-positives (242/1,528, 15.8%) and *Candida* spp. 123/1,528 (8%).

**Table 3.**
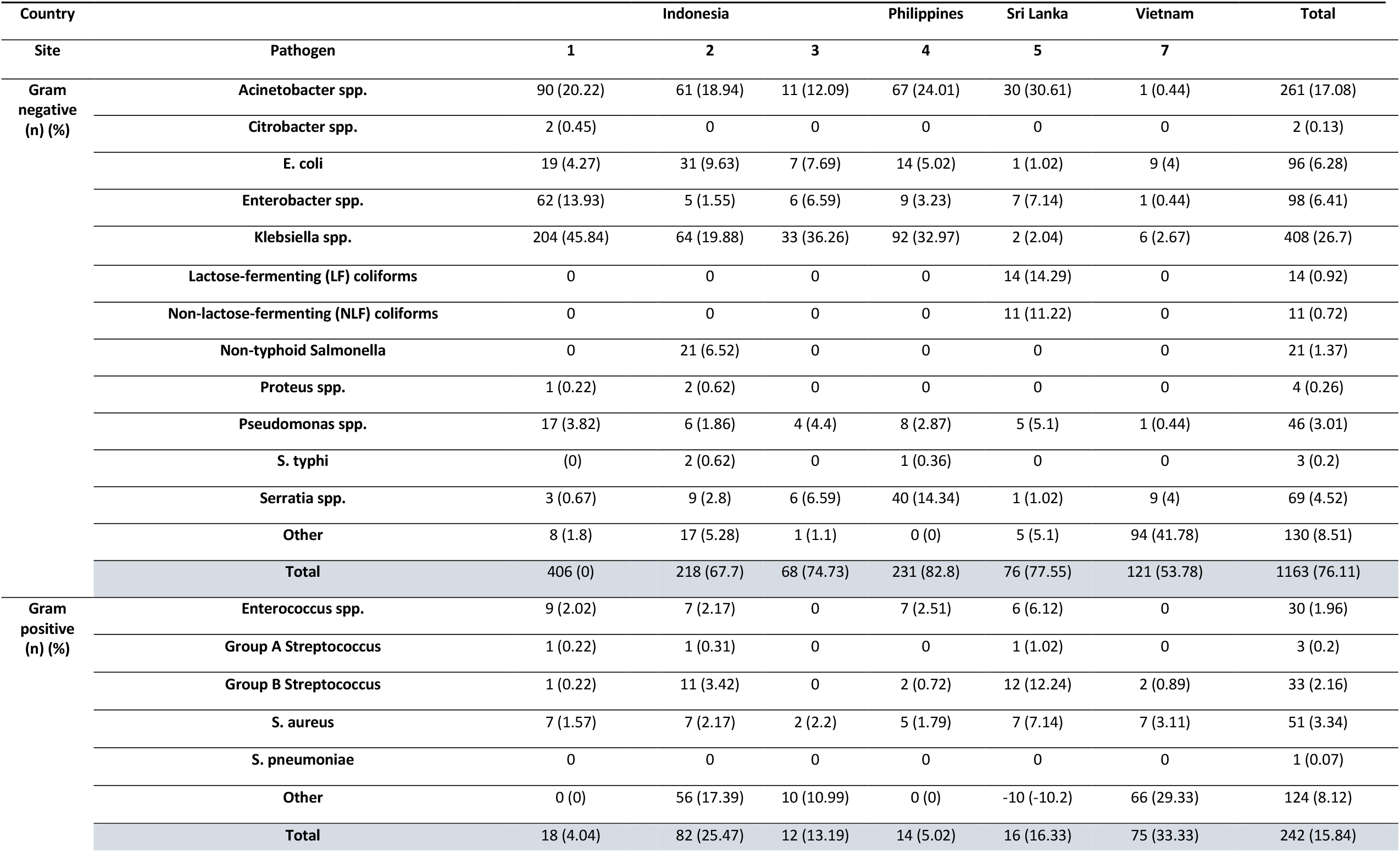
Frequency of bacterial isolates in neonatal blood cultures by site (n=1,405)

**Table 4.**
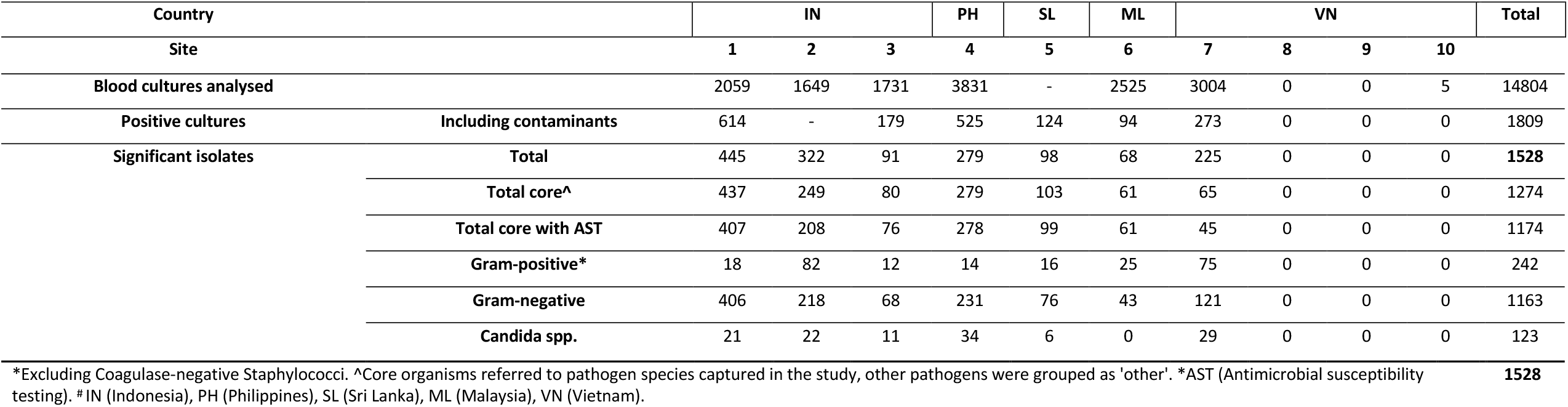
Blood culture denominators by site.

The species identified in positive cultures are outlined in Figure 2. and Table 4. The most frequently isolated pathogens were *Klebsiella* spp. (408/1,528, 26.7%), *Acinetobacter* spp. (261/1,528, 17.08%) and *Enterobacter* spp. (98/1,528, 6.41%). The gram-positive species *S. aureus* and Group B Streptococcus only accounted for 3.34% (51/1,528) and 2.16% (33/1,528) of isolates respectively. There was a significant difference between the proportion of isolates caused by *Candida* spp. between sites with (12.2% 95%CI 9.0 – 15.9) and without anti-fungal prophylaxis (6.74%, 5.4 – 8.3%) (p=0.001).

Figure 3., Table 5., Table 6., and Table 7., outline the prevalence estimates of pooled and species-level AMR among positive bacterial isolates, while Figure 4., highlights the pooled non-susceptibility to key antimicrobials. Among gram-negative Enterobacteriacae pooled resistance to ampicillin, gentamicin, third-generation cephalosporins (ceftriaxone and/or cefotaxime) (3GC) and carbapenems was 75% (193/257), 59% (393/665), 67% (441/655) and 18.6% (125/672). Resistance to gentamicin and carbapenems was higher for gram-negative non-enterobacteriaecae at 76.6% (326/282) and 69.7% (207/297). Among gram-positives, the prevalence of methicillin-resistant *S. aureus* was 26.1% (12/46), and vancomycin-resistant Enterococcus spp. was 5.88% (4/68). Across all isolates, the prevalence of 3GC and carbapenem resistance was highest in Indonesia, Philippines and Sri Lanka, and lower in Malaysia and Vietnam. *Klebsiella spp*. and *Acinetobacter* spp., the two most prevalence species, harboured the highest prevalence of resistance to 3GCs (86%, 325/376) and carbapenems (77%, 194/252) respectively.

**Figure 3.**
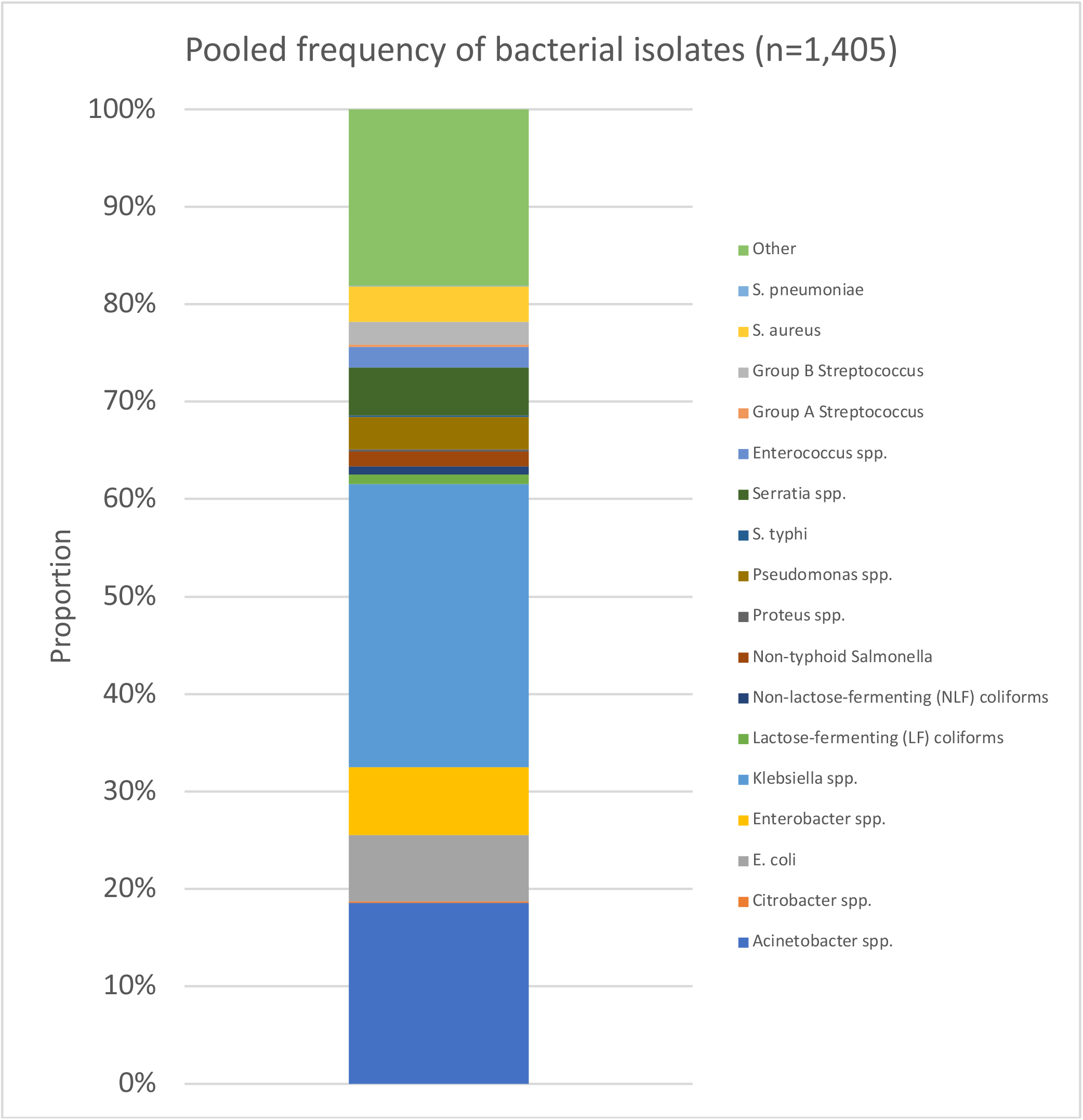
Pooled frequency of positive bacterial isolates.

**Figure 4.**
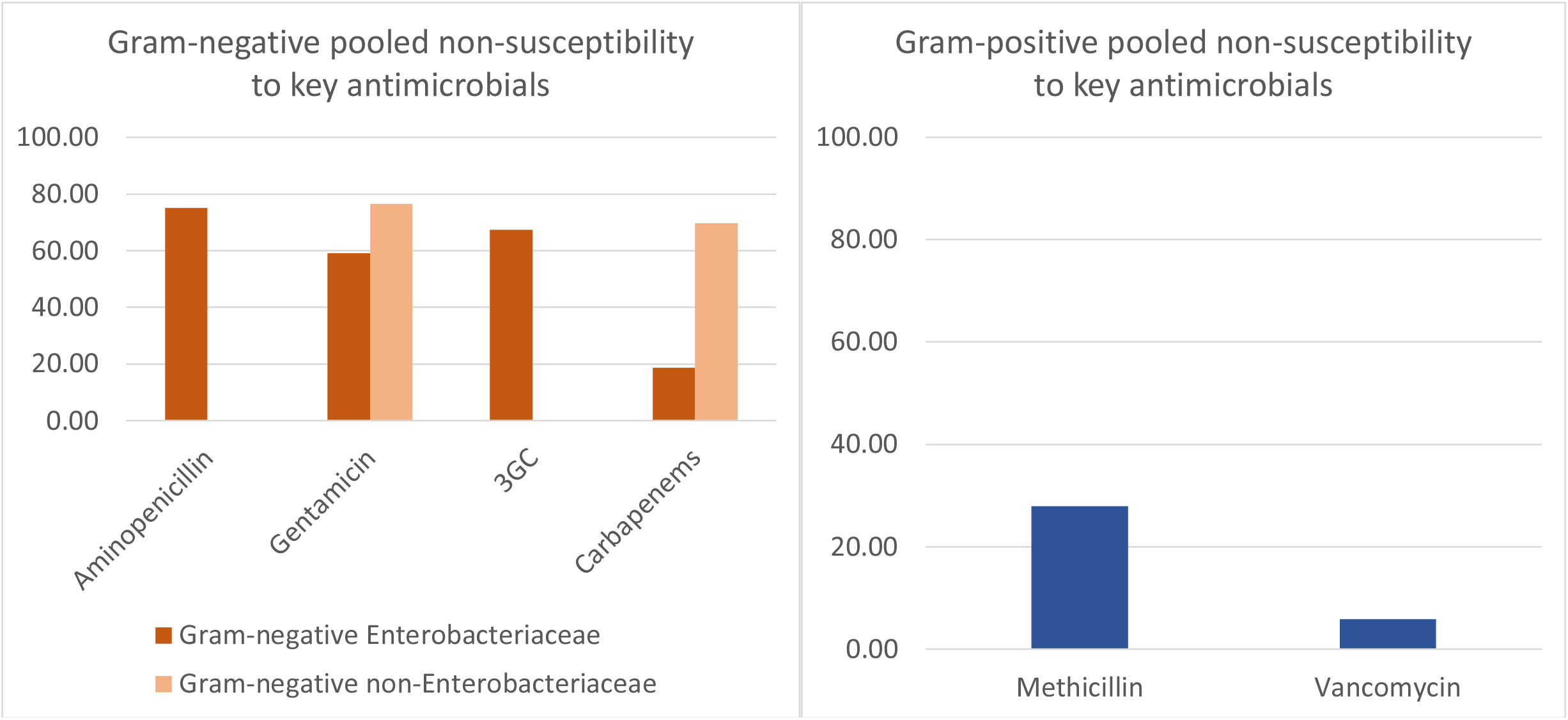
Pooled non-susceptibility to key antimicrobials for a) Gram-negative pathogens and b) Gram-positive pathogens.

**Table 5.** Pooled antimicrobial susceptibility of neonatal blood culture isolates by site and pathogen type.

**Figure 3. Pooled frequency of positive bacterial isolates**
|  |  |  |  |  |  |  |  |  |
| --- | --- | --- | --- | --- | --- | --- | --- | --- |
| Antimicrobial | Indonesia |  |  | Philippines | Sri Lanka | Malaysia | Vietnam | Total |
|  | 1 | 2 | 3 | 4 | 5 | 6 | 7 |  |
|  | (n/N) (%) | (n/N) (%) | (n/N) (%) | (n/N) (%) | (n/N) (%) | (n/N) (%) | (n/N) (%) | (n/N) (%) |
| Gram-negative non-Enterobacteriaceae |  |  |  |  |  |  |  |  |
| Amikacin | 66/97 (68.04) | 38/62 (61.29) | 8/15 (53.33) | 3/70 (4.29) | 28/34 (82.35) | 0/6 (0) | 0/1 (0) | 143/285 (50.18) |
| Carbapenems | 73/102 (71.57) | 49/62 (79.03) | 7/15 (46.67) | 49/75 (65.33) | 28/35 (80) | 0/6 (0) | 1/2 (50) | 207/297 (69.7) |
| Cefepime | 42/58 (72.41) | 60/62 (96.77) | 12/15 (80) | 54/75 (72) | 0/0 (0) | 0/5 (0) | 1/2 (50) | 169/217 (77.88) |
| Ceftazidime | 57/79 (72.15) | 58/62 (93.55) | 12/15 (80) | 53/75 (70.67) | 30/32 (93.75) | 0/6 (0) | 0/2 (0) | 210/271 (77.49) |
| Ciprofloxacin | 61/86 (70.93) | 54/62 (87.1) | 0/15 (0) | 48/75 (64) | 22/27 (81.48) | 0/6 (0) | 1/2 (50) | 186/273 (68.13) |
| Co-trimoxazole | 9/29 (31.03) | 18/57 (31.58) | 0/11 (0) | 16/67 (23.88) | 24/26 (92.31) | 0/0 (0) | 0/0 (0) | 67/190 (35.26) |
| Gentamicin | 69/92 (75) | 57/62 (91.94) | 12/15 (80) | 51/75 (68) | 25/30 (83.33) | 1/6 (16.67) | 1/2 (50) | 216/282 (76.6) |
| Piperacillin-Tazobactam | 71/100 (71) | 58/62 (93.55) | 9/15 (60) | 53/75 (70.67) | 0/0 (0) | 0/6 (0) | 0/1 (0) | 191/259 (73.75) |
| Tigecycline | 19/95 (20) | 11/62 (17.74) | 3/15 (20) | 0/0 (0) | 0/0 (0) | 0/0 (0) | 0/0 (0) | 33/172 (19.19) |
| Tobramycin | 0/0 (0) | 0/0 (0) | 0/15 (0) | 11/75 (14.67) | 0/0 (0) | 0/0 (0) | 1/2 (50) | 12/92 (13.04) |
| Gram-negative Enterobacteriaceae |  |  |  |  |  |  |  |  |
| 3GC | 196/251 (78.09) | 77/105 (73.33) | 38/52 (73.08) | 88/155 (56.77) | 29/35 (82.86) | 4/32 (12.5) | 9/25 (36) | 441/655 (67.33) |
| Amikacin | 115/257 (44.75) | 38/105 (36.19) | 12/52 (23.08) | 18/155 (11.61) | 7/31 (22.58) | 1/24 (4.17) | 0/12 (0) | 191/636 (30.03) |
| Aminopenicillin | 52/77 (67.53) | 50/52 (96.15) | 0/13 (0) | 43/63 (68.25) | 24/27 (88.89) | 24/25 (96) | 0/0 (0) | 193/257 (75.1) |
| Amoxicillin-Clavulanate | 180/274 (65.69) | 81/91 (89.01) | 38/52 (73.08) | 114/155 (73.55) | 30/33 (90.91) | 20/32 (62.5) | 19/24 (79.17) | 482/661 (72.92) |
| Carbapenems | 60/274 (21.9) | 13/105 (12.38) | 9/52 (17.31) | 23/155 (14.84) | 20/33 (60.61) | 0/28 (0) | 0/25 (0) | 125/672 (18.6) |
| Cefepime | 113/165 (68.48) | 41/91 (45.05) | 44/52 (84.62) | 84/155 (54.19) | 0/0 (0) | 3/32 (9.38) | 7/25 (28) | 292/520 (56.15) |
| Ceftazidime | 160/212 (75.47) | 80/91 (87.91) | 40/52 (76.92) | 83/155 (53.55) | 20/25 (80) | 3/32 (9.38) | 7/25 (28) | 393/592 (66.39) |
| Ciprofloxacin | 134/236 (56.78) | 55/105 (52.38) | 0/52 (0) | 40/155 (25.81) | 19/31 (61.29) | 8/32 (25) | 4/25 (16) | 260/636 (40.88) |
| Co-trimoxazole | 72/125 (57.6) | 49/105 (46.67) | 0/52 (0) | 51/155 (32.9) | 13/24 (54.17) | 12/32 (37.5) | 6/13 (46.15) | 203/506 (40.12) |
| Fosfomycin | 0/0 (0) | 0/0 (0) | 8/52 (15.38) | 0/0 (0) | 0/0 (0) | 0/0 (0) | 0/0 (0) | 8/52 (15.38) |
| Gentamicin | 205/262 (78.24) | 90/105 (85.71) | 34/52 (65.38) | 26/155 (16.77) | 21/34 (61.76) | 7/32 (21.88) | 10/25 (40) | 393/665 (59.1) |
| Netilmicin | 0/0 (0) | 0/0 (0) | 2/52 (3.85) | 0/40 (0) | 8/15 (53.33) | 0/0 (0) | 0/0 (0) | 10/107 (9.35) |
| Piperacillin-Tazobactam | 16/61 (26.23) | 3/12 (25) | 3/12 (25) | 0/49 (0) | 0/1 (0) | 1/10 (10) | 0/1 (0) | 23/146 (15.75) |
| Tigecycline | 5/263 (1.9) | 5/91 (5.49) | 20/52 (38.46) | 0/0 (0) | 0/0 (0) | 0/0 (0) | 0/0 (0) | 30/406 (7.39) |
| Tobramycin | 0/0 (0) | 0/0 (0) | 0/52 (0) | 2/141 (1.42) | 0/0 (0) | 0/0 (0) | 7/25 (28) | 9/218 (4.13) |
| Trimethoprim | 0/0 (0) | 43/91 (47.25) | 0/52 (0) | 0/0 (0) | 0/0 (0) | 0/0 (0) | 5/12 (41.67) | 48/155 (30.97) |

Note: Antibiotics excluded with less than a pooled totals of 50 isolates (Gram-negative) and 20 isolates (Gram-positive)

**Table 6.** Pooled antimicrobial susceptibility of neonatal blood culture isolates by site and pathogen type (continued)

| Gram Positive |  |  |  |  |  |  |  |  |
| --- | --- | --- | --- | --- | --- | --- | --- | --- |
| 3GC | 1/1 (100) | 0/10 (0) | / (0) | 0/2 (0) | 0/2 (0) | 0/6 (0) | 0/1 (0) | 1/22 (4.55) |
| Aminopenicillin | 3/13 (23.08) | 0/6 (0) | 0/2 (0) | 4/7 (57.14) | 0/6 (0) | 0/1 (0) | 0/0 (0) | 7/35 (20) |
| Methicillin | 0/2 (0) | 0/4 (0) | 0/2 (0) | 2/5 (40) | 3/7 (42.86) | 1/16 (6.25) | 6/7 (85.71) | 12/43 (27.91) |
| Clindamycin | 1/2 (50) | 4/14 (28.57) | 0/2 (0) | 1/7 (14.29) | 5/13 (38.46) | 5/21 (23.81) | 7/9 (77.78) | 23/68 (33.82) |
| Co-trimoxazole | 0/3 (0) | 0/4 (0) | 0/2 (0) | 0/0 (0) | 1/7 (14.29) | 1/17 (5.88) | 5/6 (83.33) | 7/39 (17.95) |
| Erythromycin | 0/2 (0) | 3/10 (30) | / (0) | 0/2 (0) | 3/8 (37.5) | 2/5 (40) | 2/2 (100) | 10/29 (34.48) |
| Penicillin | 5/9 (55.56) | 0/20 (0) | 1/2 (50) | 7/14 (50) | 2/21 (9.52) | 15/22 (68.18) | 6/9 (66.67) | 36/97 (37.11) |
| Teicoplanin | 0/13 (0) | 0/0 (0) | 0/2 (0) | 0/0 (0) | 0/0 (0) | 0/0 (0) | 0/7 (0) | 0/22 (0) |
| Vancomycin | 2/12 (16.67) | 0/10 (0) | 0/2 (0) | 2/12 (16.67) | 0/7 (0) | 0/18 (0) | 0/7 (0) | 4/68 (5.88) |

Note: Antibiotics excluded with less than a pooled totals of 50 isolates (Gram-negative) and 20 isolates (Gram-positive)

**Table 7.** Pooled antimicrobial susceptibility of common species to select antimicrobials by site.

| **Antimicrobial** | **Indonesia** |  |  | **Philippines** | **Sri Lanka** | **Malaysia** | **Vietnam** | **Total** |
| --- | --- | --- | --- | --- | --- | --- | --- | --- |
|  | **1** | **2** | **3** | **4** | **5** | **6** | **7** |  |
|  | (n/N) (%) | (n/N) (%) | (n/N) (%) | (n/N) (%) | (n/N) (%) | (n/N) (%) | (n/N) (%) | (n/N) (%) |
| **Aminopenicillin** |  |  |  |  |  |  |  |  |
| E. coli | 8/17 (47.06) | 26/26 (100) | 0/7 (0) | 10/14 (71.43) | 0/1 (0) | 14/15 (93.33) | 0 | 58/80 (72.5) |
| Enterobacter | 42/55 (76.36) | 5/5 (100) | 0 | 2/9 (22.22) | 4/4 (100) | 8/8 (100) | 0 | 61/81 (75.31) |
| Enterococcus | 2/8 (25) | 0/6 (0) | 0 | 4/7 (57.14) | 0/6 (0) | 0/1 (0) | 0 | 6/28 (21.43) |
| Serratia | 1/2 (50) | 6/6 (100) | 0/6 (0) | 31/40 (77.5) | 1/1 (100) | 1/1 (100) | 0 | 40/56 (71.43) |
| **Gentamicin** |  |  |  |  |  |  |  |  |
| Acinetobacter | 66/77 (85.71) | 55/57 (96.49) | 10/11 (90.91) | 48/67 (71.64) | 23/25 (92) | 1/1 (100) | 0/1 (0) | 203/239 (84.94) |
| E. coli | 9/17 (52.94) | 23/26 (88.46) | 4/7 (57.14) | 3/14 (21.43) | 0/1 (0) | 6/15 (40) | 6/9 (66.67) | 51/89 (57.3) |
| Enterobacter | 33/55 (60) | 0/5 (0) | 3/6 (50) | 3/9 (33.33) | 6/7 (85.71) | 0/8 (0) | 0/1 (0) | 45/91 (49.45) |
| Klebsiella | 161/185 (87.03) | 50/53 (94.34) | 26/33 (78.79) | 20/92 (21.74) | 0/2 (0) | 0/7 (0) | 4/6 (66.67) | 261/378 (69.05) |
| Pseudomonas | 3/15 (20) | 2/5 (40) | 2/4 (50) | 3/8 (37.5) | 2/5 (40) | 0/5 (0) | 1/1 (100) | 13/43 (30.23) |
| Serratia | 2/2 (100) | 3/6 (50) | 1/6 (16.67) | 0/40 (0) | 1/1 (100) | 1/1 (100) | 0/9 (0) | 8/65 (12.31) |
| **3rd Generation Cephalosporins** |  |  |  |  |  |  |  |  |
| E. coli | 9/19 (47.37) | 3/26 (11.54) | 4/7 (57.14) | 4/14 (28.57) | 0/1 (0) | 2/15 (13.33) | 5/9 (55.56) | 27/91 (29.67) |
| Enterobacter | 26/44 (59.09) | 5/5 (100) | 3/6 (50) | 2/9 (22.22) | 5/6 (83.33) | 0/8 (0) | 0/1 (0) | 41/79 (51.9) |
| GBS~ | 1/1 (100) | 0/9 (0) | 0 | 0/2 (0) | 0/2 (0) | 0/5 (0) | 0/1 (0) | 1/20 (5) |
| Klebsiella | 158/183 (86.34) | 53/53 (100) | 30/33 (90.91) | 77/92 (83.7) | 2/2 (100) | 1/7 (14.29) | 4/6 (66.67) | 325/376 (86.44) |
| Serratia | 2/2 (100) | 3/6 (50) | 1/6 (16.67) | 5/40 (12.5) | 1/1 (100) | 1/1 (100) | 0/9 (0) | 13/65 (20) |
| **Carbapenems** |  |  |  |  |  |  |  |  |
| Acinetobacter | 68/85 (80) | 47/57 (82.46) | 6/11 (54.55) | 46/67 (68.66) | 27/30 (90) | 0/1 (0) | 0/1 (0) | 194/252 (76.98) |
| E. coli | 5/17 (29.41) | 9/26 (34.62) | 2/7 (28.57) | 2/14 (14.29) | 0/1 (0) | 0/15 (0) | 0/9 (0) | 18/89 (20.22) |
| Enterobacter | 8/56 (14.29) | 0/5 (0) | 1/6 (16.67) | 0/9 (0) | 6/7 (85.71) | 0/4 (0) | 0/1 (0) | 15/88 (17.05) |
| Klebsiella | 47/195 (24.1) | 4/53 (7.55) | 6/33 (18.18) | 20/92 (21.74) | 1/2 (50) | 0/7 (0) | 0/6 (0) | 78/388 (20.1) |
| Pseudomonas | 5/17 (29.41) | 2/5 (40) | 1/4 (25) | 3/8 (37.5) | 1/5 (20) | 0/5 (0) | 1/1 (100) | 13/45 (28.89) |
| Serratia | 0/3 (0) | 0/6 (0) | 0/6 (0) | 1/40 (2.5) | 0/1 (0) | 0/1 (0) | 0/9 (0) | 1/66 (1.52) |
| **Penicillin** |  |  |  |  |  |  |  |  |
| Enterococcus | 2/7 (28.57) | 0/6 (0) | 0 | 4/7 (57.14) | 0/6 (0) | 0 | 0 | 6/26 (23.08) |
| GBS | 0 | 0/9 (0) | 0 | 0/2 (0) | 0/12 (0) | 0/5 (0) | 0/2 (0) | 0/30 (0) |
| S. aureus | 2/1 (200) | 0/4 (0) | 1/2 (50) | 3/5 (60) | 2/2 (100) | 15/16 (93.75) | 6/7 (85.71) | 29/37 (78.38) |
| **Methicillin** |  |  |  |  |  |  |  |  |
| S. aureus | 0/2 (0) | 0/4 (0) | 0/2 (0) | 2/5 (40) | 3/7 (42.86) | 1/16 (6.25) | 6/7 (85.71) | 12/43 (27.91) |
| **Vancomycin** |  |  |  |  |  |  |  |  |
| Enterococcus | 2/6 (33.33) | 0/6 (0) | 0 | 2/7 (28.57) | 0/0 (0) | 0/1 (0) | 0 | 4/20 (20) |
| S. aureus | 0/6 (0) | 0/4 (0) | 0/2 (0) | 0/5 (0) | 0/7 (0) | 0/16 (0) | 0/7 (0) | 0/47 (0) |

Note: Species with less than 15 total isolates omitted from table. ~GBS (Group B Streptococcus)

Of the 804 positive bacterial isolates from Indonesia, 710 (88%) had susceptibility data and were included in the Bayesian WISCA model. Estimated coverage was 25% (95% credible interval 22 to 29%) for aminopencillin plus gentamicin, 20% (17 to 23%) for 3GC, and 65% (62 to 69%) for carbapenems. More detailed results of these analyses have previously reported [16].

## Discussion

This study addressed important epidemiological gaps by evaluating the causes of neonatal sepsis, and the associated AMR burden in 10 sites across five countries in South and Southeast Asia. It demonstrated a predominance of gram-negative pathogens associated with alarming levels of resistance to commonly recommended empirical agents. These results highlight the urgent need for updated empirical treatment regimes, novel antimicrobial agents, and studies to understand the mechanisms of AMR development and spread in high-burden settings.

These findings align with observational studies elsewhere in Asia and Africa suggesting that gram-negative bacteria are the major cause of neonatal sepsis in resource-limited settings [6-10]. The predominance of *Klebsiella* spp. and *Acinetobacter* spp., which are typically associated with nosocomial infections, is consistent with findings elsewhere and underscores the likely role of horizontal transmission in the acquisition of AMR in these settings[8,15,17]. In contrast to high-income countries, the gram-positive bacteria Group B Streptococcus and *S. aureu*s, appear to be less clinically important. Although not captured in this study, studies in similar settings with a high burden of *Klebsiella* spp. and *Acinetobacter* spp. indicate that the pathogen incidence between early- and late-onset infections were relatively similar[8,17].

The leading sepsis pathogens in this study all demonstrated concerning levels of resistance to major antimicrobial classes. These resistance rates were similar to those previously observed [8,9,17], highlighting the urgent need for novel antimicrobial agents to effectively treat the growing number of neonates in low- and middle-income countries with multi-drug resistant infections. The current WHO empirical treatment regimens do not provide sufficient coverage for the pathogen distribution and AMR burden in these settings[18]. In response, sites adapted their local regimes to improve likelihood of treatment success, but this was associated with high use of ‘watch’ classified antimicrobials and considerable variation in regimes used. Updated empirical treatment guidelines in settings with a high burden of multidrug resistant infections are therefore required to help inform healthcare providers and harmonise global practices. This guidance will require both greater data on the genetic mechanisms of resistance, as well as pathogen distribution and resistance patterns by early- and late-onset infections to ensure they provide sufficient and appropriate coverage.

Of note, the high burden of AMR observed in this study occurred in settings with well-established IPC programs. Most sites had deployed multiple strategies to mitigate AMR transmission but faced high clinical workloads compounded by limited staffing and resources. The variable gaps in each program, and the persistence of AMR despite these efforts demonstrates the need improved data on AMR transmission routes so that targeted interventions can be implemented to reduce the spread of AMR.

An important finding in this study was the high proportion of infections caused by *Candida spp*. (8%). Whilst anti-fungal prophylaxis was implemented in two-sites, the incidence of *Candida* spp. also remained considerable (6.7%) in sites not implementing prophylaxis. Given this burden of *Candida* spp. infections and demonstrated efficacy of anti-fungal prophylaxis in vulnerable infants, its widespread adoption in the region should be strongly considered [19].

These findings should be interpreted within the context of some study limitations. Firstly, sites were predominantly large, tertiary-care hospitals with complex-patient loads selected through non-random sampling. The observed pathogen distribution and resistance profiles may therefore not be generalisable to other health-care settings in the region. Secondly, blood culture data were retrospective and collected at the hospital level rather than the individual patient level. This method may have introduced bias and limited analyses of the timing and clinical correlates of infection. Lastly, the incomplete species-level identification in Sri Lanka hindered, and may have biased pooled species- and family-level estimates.

This study nonetheless provides robust evidence that neonatal sepsis in these sites in South and Southeast Asia was caused predominantly by gram-negative pathogens with a high burden of AMR. It strengthens calls for updated empirical treatment regimes, and novel therapeutic agents to treat the increasing burden of multi-drug resistant infections. It also highlights the need for studies to better understand the acquisition and transmission AMR so that target interventions can developed.

## Data Availability

All data produced in the present study are available upon reasonable request to the authors

## Contributions

PCMW and BFRD designed the study. NDP, RA, LK, MEVU, GG, CL, HT, HNX and SMF collected study data. MS provided technical advice on the study design and interpretation. MH supported sites with data collection. BFRD and JB undertook the data analysis. BFRD and PCMW produced the first draft of the paper. All authors reviewed and approved the final draft of the paper.

## Acknowledgements

We would like to thank the Dr. Le Xuan Tuy from Tam Tri General Hospitals; Dr. Helen Malinda Kurniawan and Dr. Anisa Bela Anggraini Kolopaking from Cipto Mangunkusumo Hospital); Dr Naritha Vermasari and Dr Firstya Dyah from Dr Soetomo Hospital); Dr Djatnika Setiabudi, Dr Anggraini alam, Dr Adhi Kristianto Sugianli and Dr Filla Reviyani Suryaningrat from (Hasan Sadikin Hospital); for their contribution to this project.

